# Generative AI Use for Mental Health Support: Patterns, Correlates, and Impact among Canadian Students

**DOI:** 10.64898/2026.08.03.26359623

**Authors:** Lotenna Olisaeloka, Richard J. Munthali, Daniel V. Vigo

## Abstract

**Background:** General purpose generative AI (GenAI) chatbots are increasingly used by students for mental health support. Research on prevalence estimates vary widely, rarely link use to validated clinical measures, and have not been reported in a Canadian student population. We estimated the prevalence trends, patterns, perceived impact, and correlates of GenAI use for mental health support among Canadian university students.

**Methods:** We analysed one year (May 2025–April 2026) repeated cross-sectional data from the Canadian arm of the World Health Organization (WHO) World Mental Health International College Student survey (WMH-ICS) The primary outcome was past-year prevalence of GenAI use for mental health support. Specific use purposes, perceived impact, reasons for non-use, and future use intent were also analysed. Factors associated with GenAI use were assessed using modified Poisson regression. As a sensitivity analysis, an elastic-net penalised regression model was fitted to assess the robustness of findings to an alternative modelling approach.

**Results:** The past-year prevalence of GenAI chatbot use for mental health support was **25.2%** (95% CI 22.7–27.9), with a lifetime prevalence of **30.2%**. Use was mostly occasional and predominately for seeking mental health information, stress management, and emotional support/companionship. Regression analyses showed that students of Asian ethnicity, those with higher clinical burden/mental health diagnosis, recent adverse life experiences, weaker social support, and prior digital help-seeking behaviours were more likely to use GenAI for mental health purposes. Conversely, 2SLGBTQ+ students and those with romantic partners were less likely to seek support from GenAI chatbots. Nearly three-quarters (74.2%) of users perceived such use to have a positive impact on their mental health and emotional wellbeing. Non-users reported preference for human interaction, distrust of GenAI in mental health (67.4% each), and privacy/security concerns (50.3%). Non-use also reflected principled objections to AI, including ethical and environmental concerns, with most non-users indicating no future use intention.

**Conclusions:** GenAI chatbot use for mental health support has become commonplace among Canadian university students and is concentrated among those with greater mental health needs and fewer social support resources. Although most users perceived these tools as beneficial, their clinical effectiveness and safety remain uncertain. Rigorous prospective studies are needed to determine whether perceived benefits translate into improved mental health outcomes and whether purpose-built GenAI mental health interventions offer greater clinical benefit and safety than general-purpose chatbots.

## Background

Mental disorders are common among university students, yet most who need care do not receive it. More than one-third of students meet the diagnostic criteria for a mental disorder, and one in five experience suicidal ideation^1–3^. Despite this considerable burden, fewer than 16% receive formal treatment, with financial constraints, limited service availability, stigma, and a preference for self-management representing key barriers to care^1,4^. Against this backdrop, general-purpose generative artificial intelligence (GenAI) chatbots have become widely available, and students are increasingly using them for mental health and emotional support. These technologies were not developed or validated as therapeutic interventions, and their growing use for this purpose represents an emerging health behaviour with implications for student mental health and wellbeing.

General-purpose GenAI chatbots are large language or multi-modal conversational agents that execute common tasks like question answering, summarization, coding, and brainstorming, by generating dynamic responses based on prior training data. These chatbots such ChatGPT, Gemini, and Claude have become the most rapidly adopted technology in decades. For instance, 66% of Canadians reported using GenAI chatbots in 2025. Young people are among the most frequent users, primarily engaging with GenAI for study, work and leisure^5^. They are also increasingly turning to AI to manage their mental health and emotional needs especially given the barriers they face in accessing formal care^6,7^. However, the extent, patterns and impact of this use remains poorly characterized. This gap is important because GenAI tools are prone to hallucination and sycophancy, with emerging research revealing that they can provide wrong information, respond inappropriately to mental health disclosures, express stigma, or fail in safety-critical exchanges. Furthermore, interactions with general-purpose chatbots have been linked to serious harms, including psychosis and suicide, promoting increased research focus and regulatory attention^8,9^.

What is known about GenAI use for mental health support is still thin, and the estimates vary widely with population and question wording. An early US study among 428 university students conducted about one year post-ChatGPT release, found that while 49% had used AI chatbots, only 5% reported mental health related use cases^10^. More recent studies show use figures ranging from 13.1% among US adolescents to 35% of the general adult population seeking mental health advice from an AI chatbot^11,12^. The existing literature is predominantly constrained by online, convenience-sampling designs, which may fail to generate reliable prevalence estimates. A recent review highlighted that many such AI use surveys do not adhere to rigorous scientific standards or basic reporting methodologies^13^. Furthermore, most studies relied on a single timepoint, thus were unable to capture how usage patterns evolved over time. Existing research also rarely examined the correlates of GenAI use or linked usage to validated clinical measures and theoretically informed variables. For instance, the largest representative survey to date omitted any measurement of diagnosed mental illness, leaving it unclear whether GenAI use aligns with clinical need. Additionally, few studies have characterized the specific patterns and purposes of use, and almost none have examined non-use, including reasons for abstaining and willingness to use, despite growing interest in principled non-use and questions about whether AI adoption is equitable or appropriate. Lastly, to our knowledge, no published study has examined this emerging health behaviour in a Canadian student population, or utilized the World Mental Health International College Student (WMH-ICS) survey instrument.

We addressed these critical gaps using one-year trend data from the World Health Organization (WHO) WMH-ICS survey at a major Canadian university. Our specific objectives were to:

i. Estimate the lifetime and past-year prevalence of GenAI chatbot use for mental health support and describe how these rates trended across the study period.
ii. Characterize the distinct patterns and specific purposes of GenAI chatbot use;
iii. Identify the reasons for non-use and future utilization intentions among never-users;
iv. Describe the perceived impact of GenAI chatbot use for mental health and emotional support; and
v. Examine the sociodemographic and clinical correlates of use

Ultimately, these timely empirical data will help understand how emerging AI technologies are influencing student help-seeking behaviours and mental wellbeing. Findings will inform clinical, governance, and policy efforts to safeguard student health and wellbeing whilst navigating the unprecedented digital help-seeking landscape.

## Methods

### Study Design and Data Source

The study utilized data from the Canadian arm of the World Health Organization (WHO) World Mental Health International College Student (WMH-ICS) survey^14^. The WMH-ICS initiative was established to generate global epidemiological data on the prevalence, risk factors and treatment patterns of mental disorders and to help develop evidence-based interventions^15^. The survey is administered as a self-completed, web-based questionnaire, adapted from the WHO Composite International Diagnostic Interview (CIDI), to estimate the current, lifetime and 12-month prevalence of mental and substance use disorders. It also collects rich contextual information on sociodemographic, risk and protective factors, symptom severity, help-seeking behaviour, treatment history, and perceived barriers to care^15^. Since its inception, the WMH-ICS has been completed by over 100,000 students across 18 countries^14,16^.

This study analyzed 12 months (May 2025 to April 2026) of repeated cross-sectional survey from the WMH-ICS deployment at the University of British Columbia (UBC) in Canada. The survey invites biweekly stratified random samples of enrolled students, drawn to be representative of the broader student population, with stratification by sex, age group, student level/year of study, and international-student status. Recruitment follows a rigorous multi-contact protocol (including initial email invitation, two reminder emails, and telephone follow-up of non-respondents), described in detail in the WMH-ICS Canada survey protocol^14^. The survey’s adjusted response rate is 43.04% using the American Association for Public Opinion Research Response Rate 1 weighted (RR1w) method for two-phase designs, accounting for the initial survey response rate (27.9%) and the second-phase follow-up response rate among non-respondents (24.4% for those with phone numbers on file, and 5.4% for those without)^17^.

The survey has been ongoing at UBC since 2020. Items on general-purpose GenAI use for mental health and emotional wellbeing were added to the treatment-seeking module in April 2025 following an ethics amendment. The analytic sample comprised all respondents with a valid response to the GenAI use item. Further details on the survey methodology can be assessed from the published survey protocol paper^14^.

### Ethics

All procedures contributing to this study adhered to the ethical standards of the relevant national and institutional committees on human experimentation and with the Helsinki Declaration of 1975, as revised in 2008. All procedures involving human subjects were approved by the Behavioural Research Ethics Board of the University of British Columbia (H19-02538). Participation was entirely voluntary and students provided informed consent prior to completing the survey. Upon completion, all students were provided a tailored list of support services based on their survey responses^14^.

### Study Variables

#### Generative AI use for mental health

The primary outcome was past-year (12-month) GenAI chatbot use for mental health or emotional support. Lifetime use (ever-use) was also reported descriptively a secondary outcome. Users answered additional questions on purpose and perceived impact of use, while never-users were routed to the willingness to use item. **Supplementary Table 1** contains the GenAI questionnaire items added to the WMH-ICS survey.

#### Correlates of GenAI Use

Candidate correlates were selected on theoretical grounds, guided by predisposing, enabling, and need domains from the Andersen’s Behavioural Model of Health Service Use^18^. Predisposing variables comprised age group (ordinal bands: 16–20, 21–24, 25– 29, 30, 34, ≥35), gender (Man/Woman / Non-binary, two-spirit & other), race/ethnicity, (White / Asian / Black / Hispanic / First Nations, Inuit, Metis / Other including multi-racial), sexual identity (Heterosexual / Lesbian, gay, bisexual / Other including asexual), relationship status (Married / Dating or in a relationship / Single), immigration indicator (Born in Canada vs outside) and international student status (Yes / No). Enabling variables comprised prior use of internet or digital tools for mental health support, willingness to use digital mental health tools, lifetime counselling, and 12-month medication or psychological treatment status. Need was operationalised clinically through validated DSM-V based 12-month screens for common and severe mental disorders, substance-use disorders, and suicidality, and through self-rated mental health, psychosocial stressors (including academic and financial stress) and any recent adverse life experience. Social connection as a need indicator was captured through online and in-person contact with friends or relatives, perceived social support (reliance on others for support) and support-network size (no of people participants could rely on).

### Statistical Analysis

Categorical variables were summarised as n (%) and continuous variables as mean [SD] or median [IQR] if skewed. A descriptive table presented participant characteristics overall and by past-year use. Past year and lifetime prevalence were estimated with a 95% Wilson score confidence interval [CI]. Patterns of use (frequency and purpose), reasons for non-use, perceived impact, and future intent were summarised descriptively and with figures. Free-text responses for non-use were analysed by inductive reflexive thematic analysis.

Correlates of past-year use were examined using modified Poisson regression, with robust (sandwich) standard errors. The modified Poisson estimator returns a directly interpretable prevalence ratios (PR) and avoids overstating associations like odds ratios often do when the outcome is common^19^. To ensure unbiased estimation, all models were adjusted for the sample-design variables (survey week, sex, and year of study). Sociodemographic variables and other theory-informed enabling variables including prior use and willingness to use digital tools for mental health purposes were retained a priori. For Clinical need, the individual disorders were collapsed as typically done in psychiatric epidemiology to avoid signal-splitting across correlated screens. This resulted in the clinical burden index: a linear count (0/1/2/3+) of past-year disorder screens and suicidality, capturing cumulative psychiatric morbidity. Multicollinearity was assessed by VIF (flag > 5) and model discrimination by the area under the ROC curve (AUC). Missing data for the final model totalled 5.3% with maximum single-variable missingness of 2.3%, Missingness was unrelated to the outcome (5.9% of users vs. 5.1% of non-users, p = 0.72) and jointly unpredicted by covariates (likelihood-ratio p = 0.99), supporting complete-case analysis as the primary approach. Seven interaction terms specified a priori were tested. As a sensitivity analysis, an elastic-net penalised regression was fitted to assess whether the model’s associations were robust to data-driven variable selection. Two-sided p < 0.05 was used for interpreting the final model. Analyses were conducted in R (version 4.3.1) with packages *dplyr* and *haven* for data management; *sandwich* and *lmtest* for robust SE; *car* for multicollinearity; *pROC* for model discrimination; *glmnet* for elastic net sensitivity analysis; and *ggplot2* for figures^20^.

## Results

### Sample characteristics

Of 1,067 students who answered the GenAI module, 269 (25.2%) reported using a general-purpose GenAI chatbot for mental health support in the past 12 months. The sample carried a high symptom burden: 41.5% rated their mental health as fair or poor, 21.8% screened positive for a 12-month major depressive episode, 31.7% for PTSD, and 28.9% reported past-year suicidal ideation or attempt. Full characteristics, overall and by past-year use, are shown in **Table 1**. On bivariate inspection, few sociodemographic variables such as age, race and relationship status differed significantly between GenAI users and non-users, with stronger statistical differences observed across three coherent domains: clinical and stress-related need, digital help-seeking behaviour, and social connection. GenAI users had higher rates of depression, social anxiety, PTSD, eating disorder, severe mental illness and psychosocial stress compare to non-users (all p<0.001). Users were more likely to have previously sought mental health help online (p<0.001) and to report willingness to use digital mental health resources. Compared to non-users, students who use GenAI for mental health or emotional support reported lower in-person social contact and number of people they could rely on for support or comfort (p < 0.001).

**Table 1.**
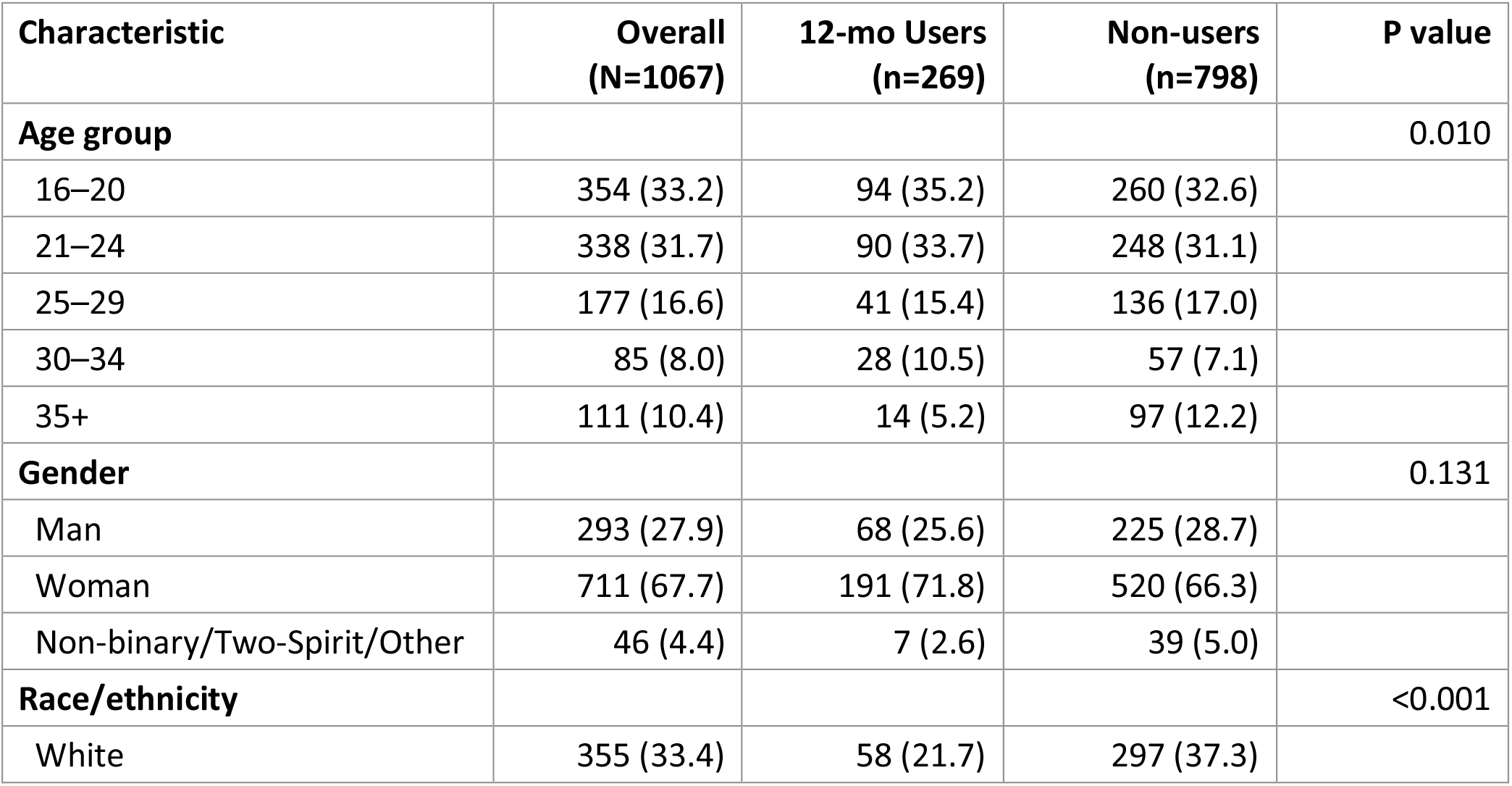
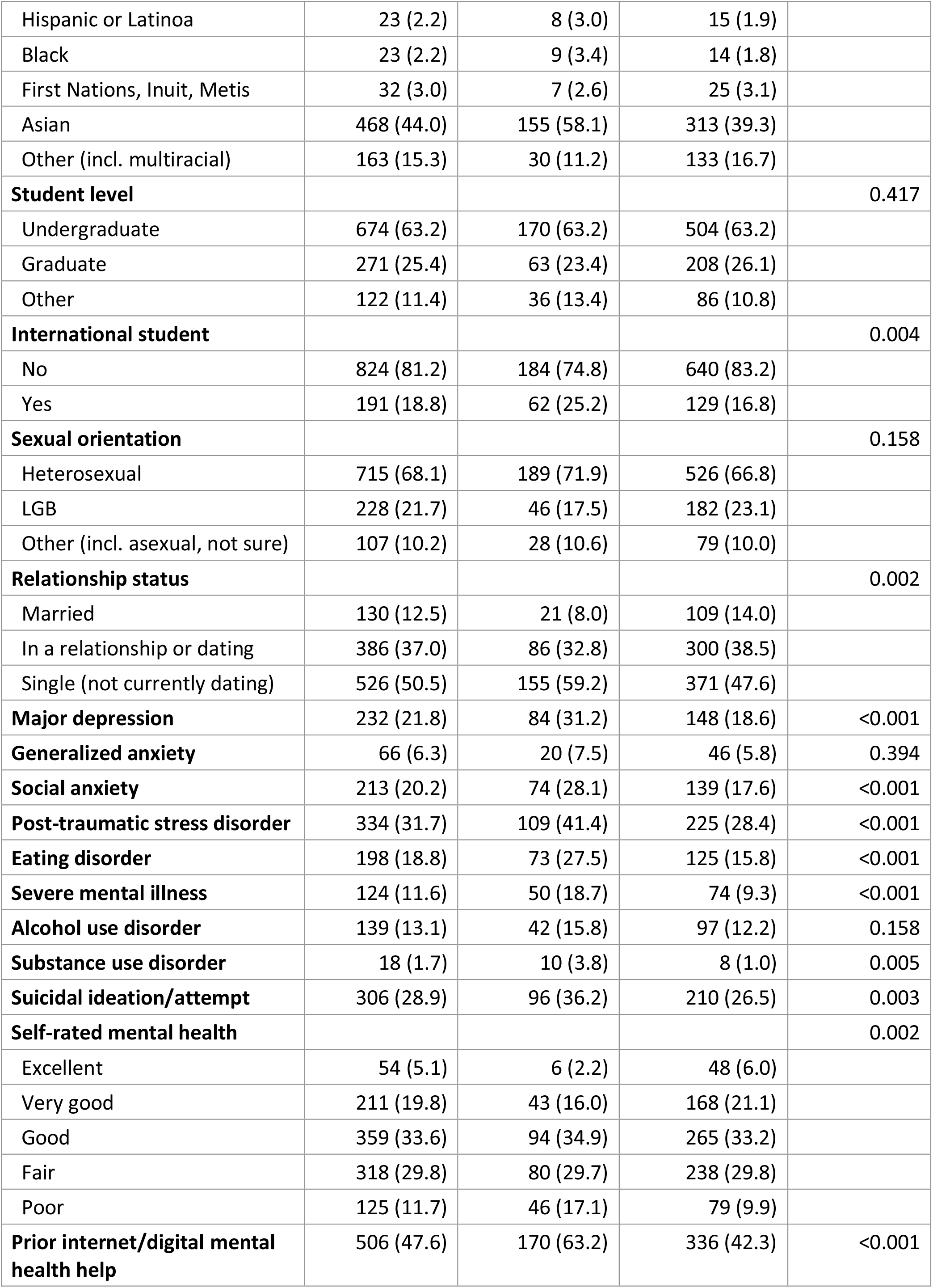
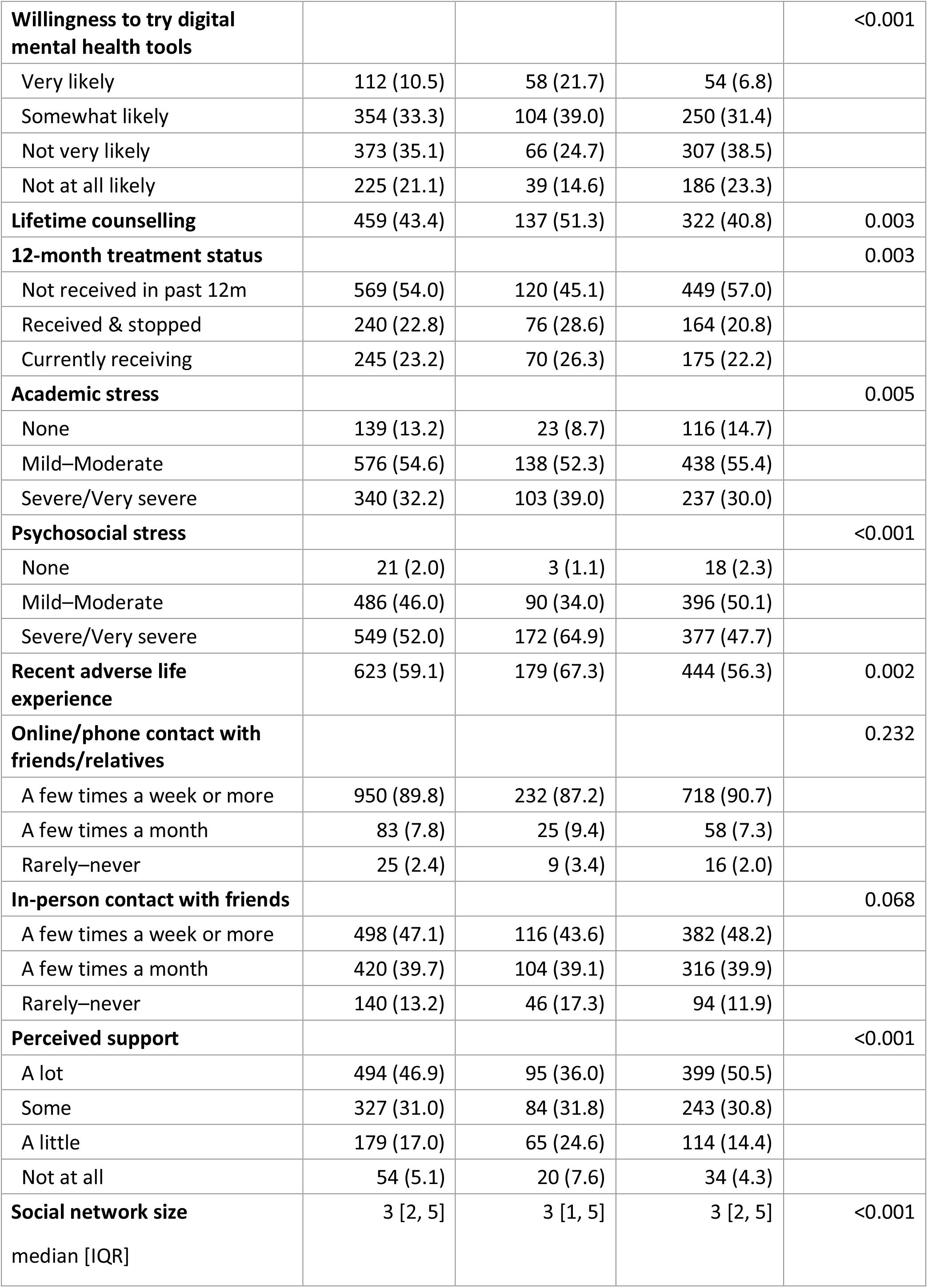
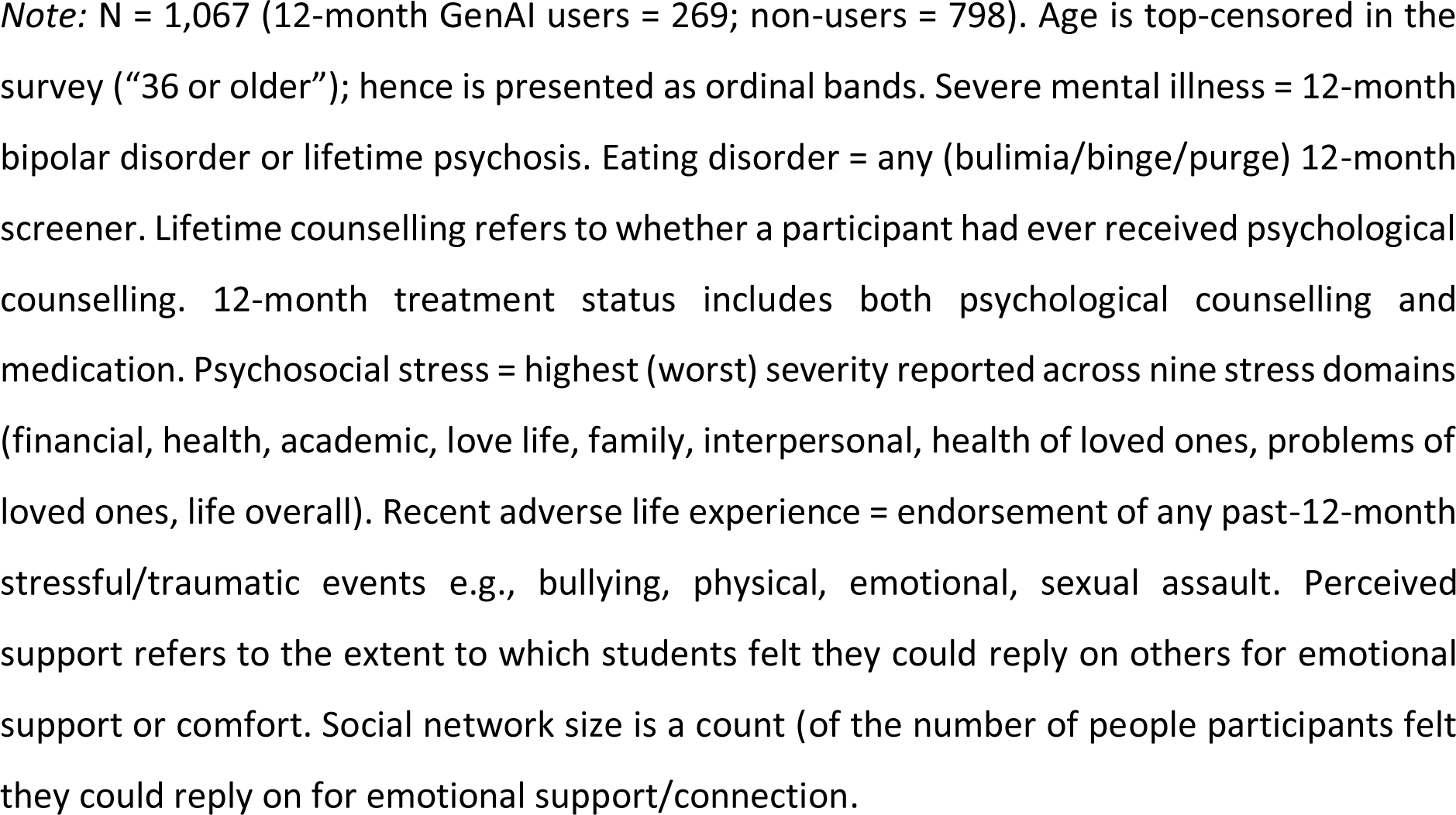
Descriptive characteristics of the analytic sample, overall and by 12-month GenAI use for mental health and emotional support.

### Prevalence and trends of GenAI use for mental health and emotional support

Past-year prevalence of GenAI use for mental health or emotional support was 25.2% [95% CI: 22.7–27.9], and lifetime (ever) use was 30.6% [95% CI: 28.0–33.5%). **Figure 1a** shows that the monthly prevalence estimates fluctuated within a relatively narrow band across the observed 12-month study period. Among the participants who utilized GenAI chatbots, use was predominantly occasional (less than monthly) rather than habitual as shown in **Figure 1b**.

**Figure 1a:**
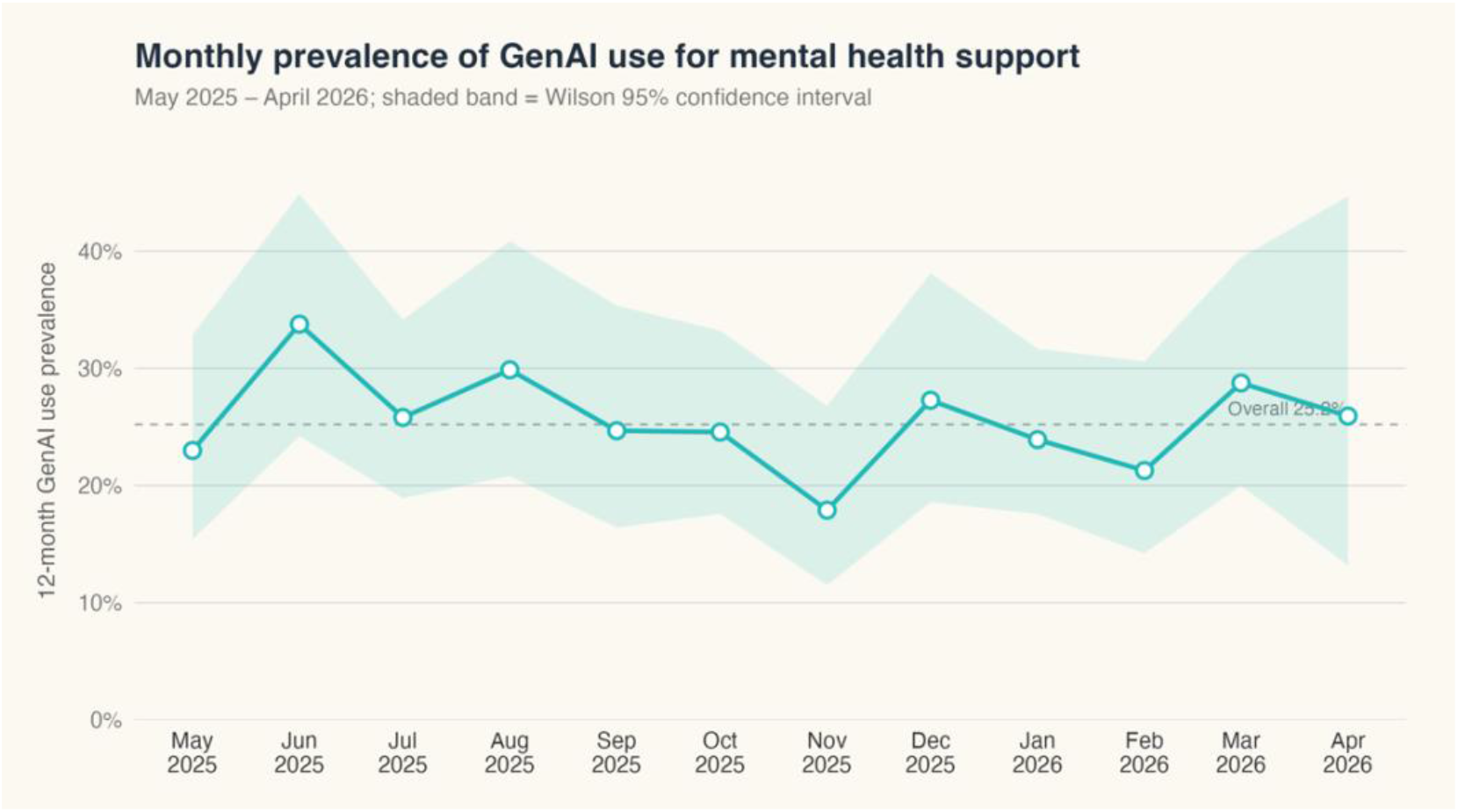
One year prevalence trend of general purpose GenAI Chatbot (e.g., ChatGPT, Gemini, Claude, DeepSeek) use for mental health and emotional support purposes.

**Figure 1b:**
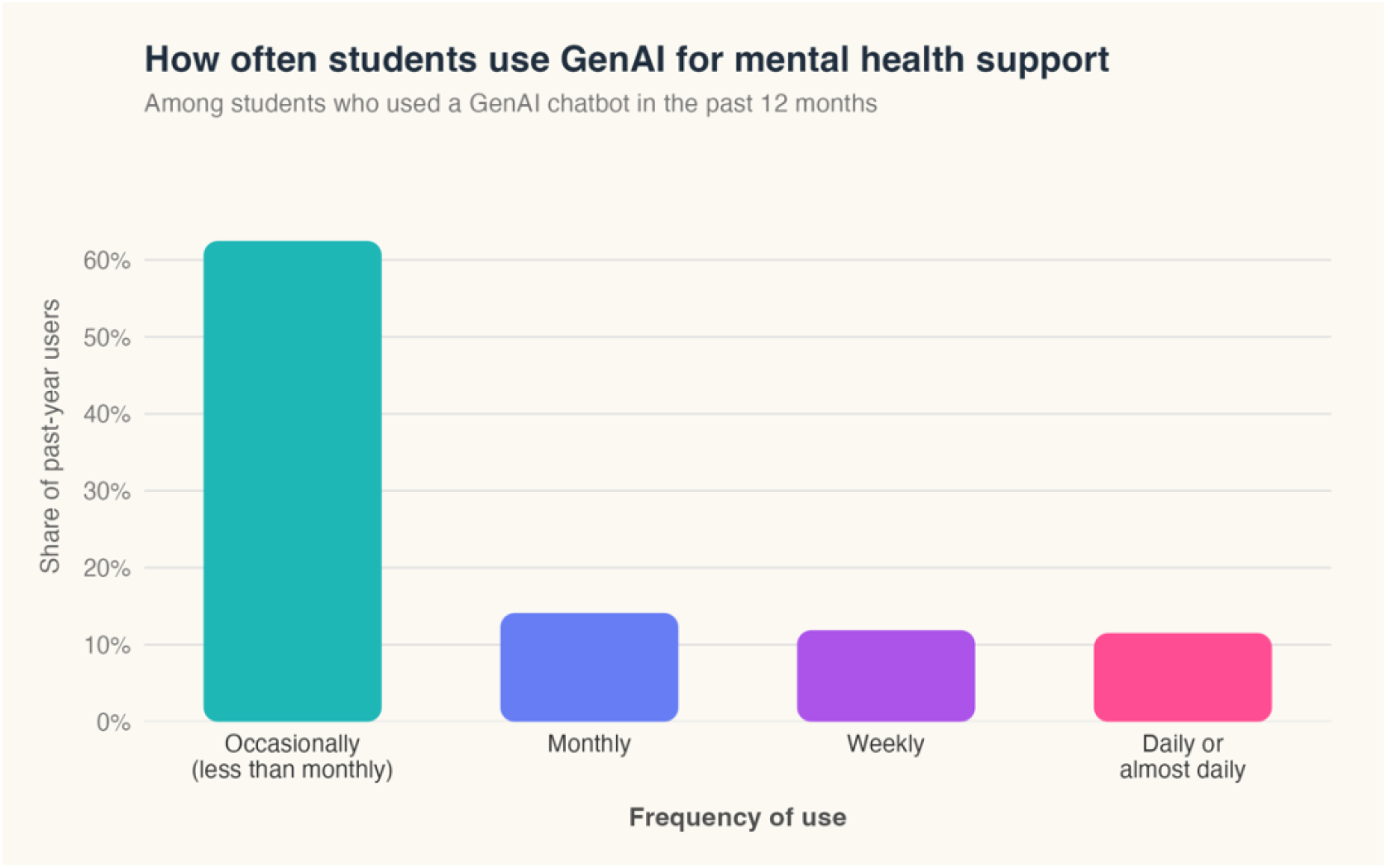
Frequency of GenAI chatbot use for mental health and emotional support purposes among Canadian students.

### Usage patterns and reasons for use and non-use

The most common use purposes were obtaining information or advice about mental health (58.3%) and managing stress (57.4%), followed by emotional support or companionship (46.9%), general well-being (38.0%), and use as an alternative or supplement to therapy (34.6%). Advice about substance use was an uncommon use case (8.0%) (**Figure 2**). Multiple-purpose use was the norm as users selected a median of 2 purposes and 67% reported using GenAI for two or more distinct purposes. The most common pairing was information-seeking combined with stress management (n = 108), followed by stress management with emotional support (n = 98) and information-seeking with emotional support (n = 86). The thematic analysis of 22 free-text responses pointed mainly to venting and GenAI use as an active “sounding-board” for reasoning through problems, processing emotions, or seeking reassurance, including in acute distress when a human source was unavailable.

**Figure 2:**
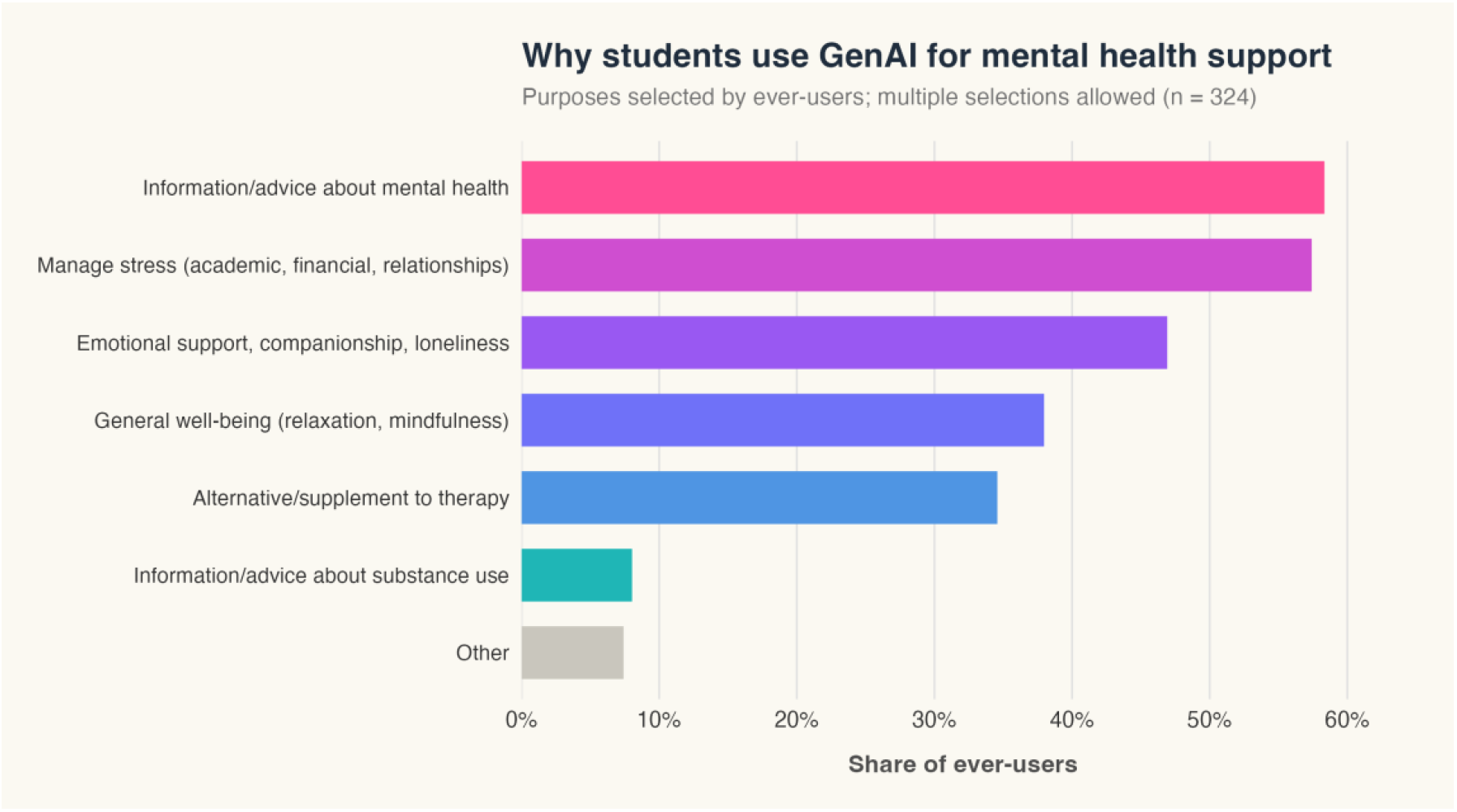
Purposes/use cases for GenAI chatbots in managing students’ mental health and emotional needs*. Individual bars represent survey options; because the survey item was multi-select, percentages are of respondents (not responses) and do not sum to 100%*.

Among the 731 non-users, the two dominant reasons were preferring human interaction and distrust of AI to handle mental health, each cited by 67.4% of respondents. About 80% cited at least one of these predominant reasons and 56% cited both, indicating a coherent reluctance to delegate mental health support to AI (**Figure 3a**). Privacy/security was another important reason, selected by half (50.3%) of the non-users. Notably, the least common reason was being unsure how to use the tools (11.8%), pointing to non-use being primarily a deliberate preference more than a skills or awareness gap. Free-text responses (n=82) analysed thematically showed a majority of answers expressed general opposition or values-based objections to AI more than specific judgements about GenAI’s usefulness for mental health. These included ethical and moral opposition to AI, environmental reasons, and distrust of AI’s safety or accuracy (**Figure 3b**). Smaller groups cited structural or need-based reasons including already having sufficient support and having no mental-health needs. **Supplementary Table 2** presents a detailed summary of themes with illustrative quotes.

**Figure 3a:**
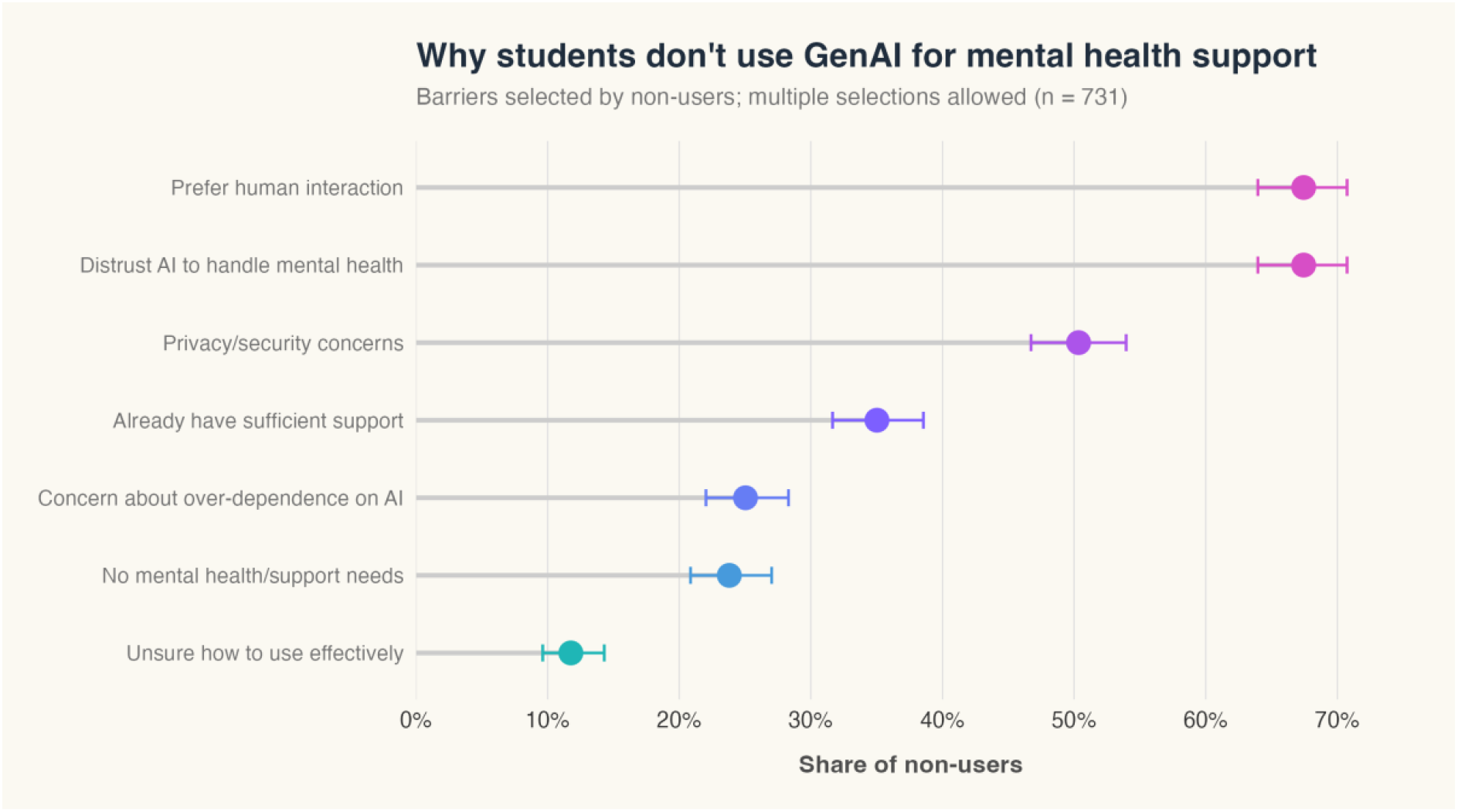
Reasons for non-use of GenAI chatbots for mental health and emotional support purposes

**Figure 3b:**
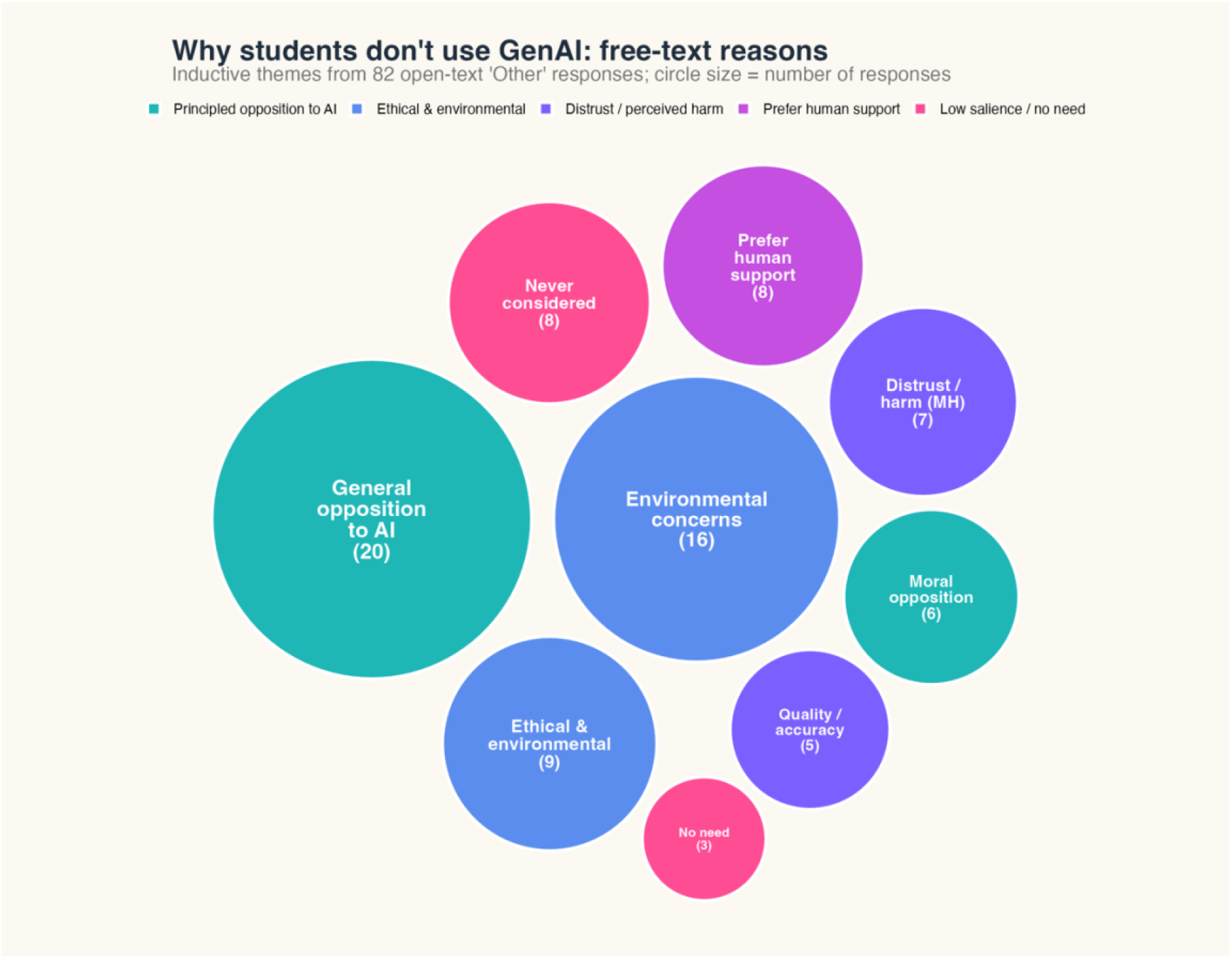
Bubble plot illustrating thematic summary of free text responses on reasons why students choose not to utilize GenAI for mental health. Bubble size represents the relative importance of various reasons with number of participants citing such reason denoted in brackets.

Of the 740 never-users, prospective intent was low: 615 (83.1%) reported being unlikely to use a GenAI chatbot if they needed support in future (63.2% very unlikely), 96 (13.0%) were neutral, and only 29 (3.9%) were likely or very likely.

### Perceived impact of GenAI Chatbot Use on mental health and emotional needs

Among the 325 ever-users who rated overall impact, 74.2% reported a positive impact (somewhat or very positive), 20.6% reported no impact, and only 5.2% reported a negative impact. Perceived positive impact was strikingly uniform across demographic and clinical subgroups, ranging roughly 68–80% in almost every stratum with overlapping confidence intervals. No demographic characteristic (age, gender, race/ethnicity, sexual orientation, student level, international status) or clinical characteristic (any of the screened disorders, severe mental illness, suicidality, treatment status) was associated with a materially different likelihood of rating GenAI as helpful (**Figure 4; Supplementary Table 3**).

**Figure 4:**
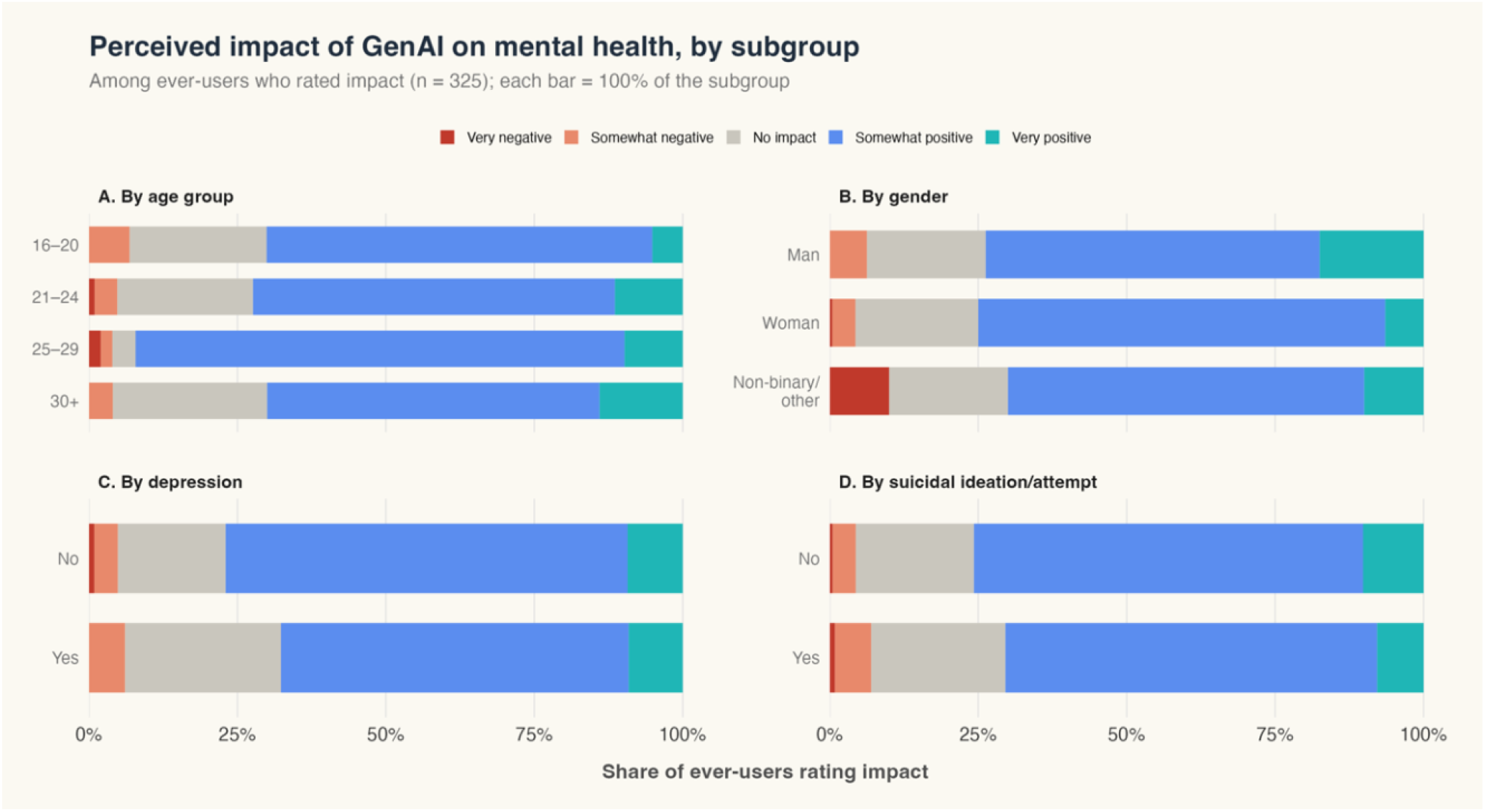
Impact of GenAI chatbots on mental health and emotional support needs. Panels show the distribution of perceived impact within each subgroup.

### Correlates of past-year GenAI use for mental health and emotional support

Among predisposing characteristics, Asian ethnicity showed the highest GenAI use prevalence relative to White students (PR 1.72, 95% CI 1.27–2.33). Conversely, being partnered or dating (0.71, 0.57–0.90), identifying as LGB (0.68, 0.51–0.91), and having social support (0.79, 0.64– 0.97) were each associated with lower use [**Table 2**]. Digital help-seeking orientation was the dominant axis. Prior use of digital tools for mental health (1.46, 1.14–1.87) and willingness to use digital mental health resources (PR 1.67, 95% CI 1.35–2.08) were the strongest correlates. Any past-year mental health treatment (medication or psychotherapy) was also associated with higher use (PR 1.27, 1.01–1.60). Clinical need operated as a cumulative gradient as each additional clinical burden indicator raised prevalence by approximately 20% (1.20, 1.08–1.32). Recent adverse life experience was also associated with higher GenAI use for mental health (PR 1.34, 1.06–1.68).

**Table 2.**
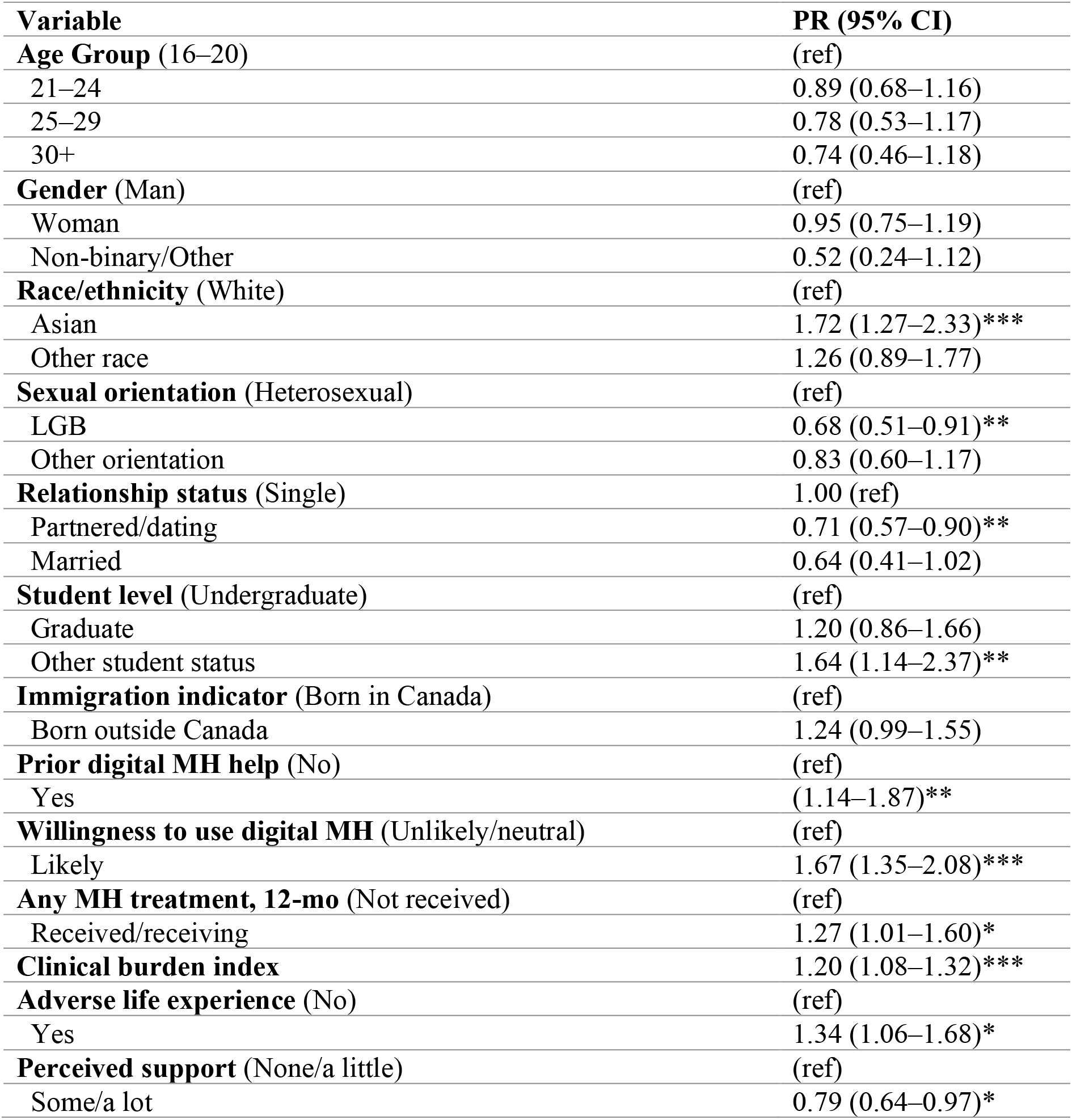
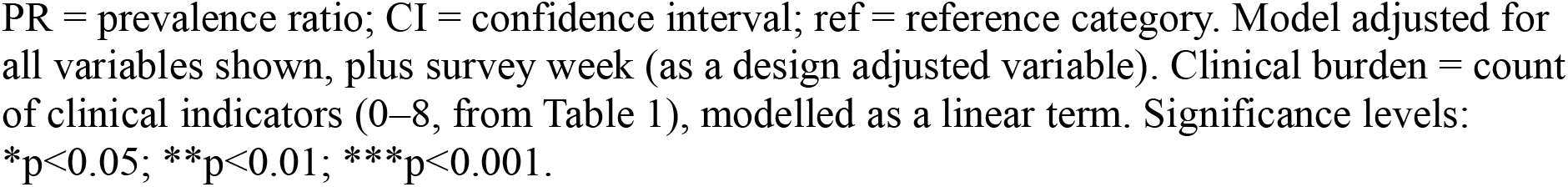
Correlates of past-year GenAI use for mental health support among Canadian university students (modified Poisson regression, n=1,010)

Model discrimination was acceptable-to-good (AUC 0.752) and the model explained a modest share of outcome variation (McFadden R²=0.10), as is typical for correlates models of individual health behaviours. Maximum VIF was 1.57, indicating no problematic collinearity. All seven pre-specified interactions were non-significant (**Supplementary Table S4**). An elastic-net penalised model retained the same core correlates and shrank to zero the variables dropped from the primary model, corroborating that the associations reflect real structure rather than artefacts of variable selection (**Supplementary Table S5**).

## Discussion

This study provides the first population-based prevalence estimates of general-purpose GenAI chatbot use for mental health and emotional support among Canadian university students. It extends previous work in other contexts by leveraging a probability-based comprehensive mental health survey which links GenAI use to validated psychiatric screeners, behavioural, clinical and social correlates.

Several principal findings emerged. First, one in four students reported using a GenAI chatbot for mental health support in the past year and more than thirty percent reported lifetime use, indicating that this health behaviour has become commonplace among this population. Second, use appeared to reflect clinical need and psychosocial vulnerability rather than sociodemographic characteristics, being concentrated among students with greater psychiatric morbidity, prior digital help-seeking, recent adverse experiences, and weaker social support. Third, students primarily used GenAI as a source of mental health information, stress management, emotional support and companionship rather than as a substitute for formal mental healthcare. Fourth, despite persistent concerns regarding the safety and reliability of these technologies, nearly three-quarters of users perceived their experiences as beneficial, with perceived helpfulness remaining remarkably consistent across demographic and clinical subgroups. Finally, non-use was largely a deliberate choice, driven by ethical and environmental concerns, preferences for human interaction, trust and privacy concerns rather than by lack of awareness or digital literacy. Collectively, these findings suggest that GenAI chatbots are now embedded within the informal student mental health support ecosystem, highlighting the need to better understand their impact and the implications for safety and governance.

Our 12-month prevalence estimate place GenAI use for mental health support at the upper end of the emerging literature, higher than both the 5% found in an early survey of US students and more recent surveys which show 13.1% prevalence among adolescents and young adults^10,11^. Although methodological differences, including survey wording, sampling strategies and study timing, preclude direct comparison, our estimate suggests that GenAI is becoming an established component of students’ help-seeking behaviour. Our findings are consistent with a Canadian opinion poll which found that nearly one in ten adults have used AI tools for mental health support, with uptake highest among younger adults^7^. Nevertheless, our sample carried a high burden of mental health symptoms, which may partly explain the higher prevalence observed relative to other studies.

Our findings provide useful insight into how students are using these technologies. Rather than replacing psychotherapy or formal treatment sources, GenAI was predominantly used for seeking mental health information, emotional support, stress management, and companionship. Coupled with the predominantly occasional frequency of use, these findings suggest that students are drawing on conversational AI episodically during periods of stress or uncertainty rather than relying on it as a substitute for ongoing mental healthcare. Taken together, these findings suggest that conversational GenAI occupies a space between traditional digital self-help resources and formal mental healthcare, providing immediate, relational, and personalized support, particularly when human-delivered care is unavailable or difficult to access. Our findings also suggest that GenAI use reflects an existing digital help-seeking orientation rather than a fundamentally new behaviour. Prior digital help-seeking and willingness to use digital mental health resources were the strongest independent correlates of use, exceeding the magnitude of most demographic and clinical factors. Rather than representing a new mode of help-seeking, conversational GenAI appears to be the latest evolution of the digital help-seeking ecosystem, which already includes internet searches, social media, and digital mental health interventions. However, the generative and personalized nature of its outputs introduce unique risks such as hallucinations, inaccurate information, anthropomorphization, and dependence^8^.

Beyond digital help-seeking orientation, clinical need emerged as another major determinant of GenAI use, with important implications for the safe deployment of these technologies. Clinical burden showed a clear gradient, with each additional clinical indicator (including major depression, social anxiety, eating disorders, bipolar disorder, psychosis, and substance use) associated with approximately a 20% higher prevalence of use. To our knowledge, this is among the first studies to demonstrate this relationship because previous surveys have rarely incorporated validated diagnostic measures. One interpretation is that students experiencing greater psychological distress are turning to GenAI to meet needs that remain unmet by existing systems of care. This finding sharpens ongoing safety concerns because the students most likely to rely on these largely unvalidated technologies are also those with the greatest clinical vulnerability, and therefore stand to experience the greatest harm if responses are inaccurate, inappropriate, or unsafe. As universities and healthcare systems consider the role of GenAI, ensuring the safety of these high-need users should therefore become an important priority.

Social context also appeared to influence GenAI use. Students who were partnered and those reporting greater perceived social support were less likely to use GenAI for mental health support. Together with the finding that nearly half of users sought emotional support or companionship, this suggests that GenAI may partially fulfil interpersonal support needs, particularly among students with fewer existing social resources. By contrast, few sociodemographic characteristics were independently associated with use. Sexual minority (LGB) students reported lower GenAI use, contrasting with previous Canadian survey findings and broader digital mental health literature, but potentially reflecting greater privacy or trust concerns or preferences for other sources of support^7^. By contrast, few sociodemographic characteristics were independently associated with use. Sexual minority (LGB) students reported lower GenAI use, contrasting with previous Canadian survey findings and broader digital mental health literature, but potentially reflecting greater privacy or trust concerns or preferences for other sources of support. Asian students were more likely than White students to report GenAI use, consistent with higher use observed among Asian students is consistent with evidence from a Canadian opinion poll which found higher utilization among ethnic minorities^7^. evidence that Asian students are less likely to access formal mental health services because of stigma, family expectations, disclosure concerns, and preferences for managing distress privately. In this context, the privacy and anonymity afforded by GenAI may make it a more acceptable source of support.²¹ Future research should examine how cultural factors shape AI-mediated help-seeking and whether GenAI mitigates or reinforces existing mental health inequities, particularly given concerns about algorithmic bias.

Compared with White students, Asian students were more likely to report using GenAI for mental health support. Although students in the heterogeneous “Other” ethnicity category also had relatively higher prevalence, this association was not statistically significant. Combining multiple distinct ethnocultural groups due to small category numbers may have obscured meaningful group-specific differences. The higher use observed among Asian students is consistent with evidence from a Canadian opinion poll which found higher utilization among ethnic minorities^7^. It may also reflect evidence from the Canadian World Mental Health International College Student (WMH-ICS) survey demonstrating lower use of formal mental health services among Asian students, often attributed to stigma, family expectations, concerns about disclosure, and preferences for managing distress privately. Within this context, GenAI may represent an acceptable avenue for support because it offers privacy and anonymity^21^. Future research should examine how cultural factors influence AI-mediated help-seeking, and whether GenAI reduces or reinforces existing mental health inequities amid concerns about algorithmic bias.

Perhaps the most distinctive contribution of this study concerns non-use. The dominant reasons for non-use were preferences for human interaction, distrust of AI, and concerns regarding privacy, ethics and security, whereas relatively few students reported uncertainty about how to use these technologies. Free-text responses reinforced this interpretation, with many participants expressing objections to AI itself, including concerns regarding environmental impacts and broader ethical issues, rather than questioning its usefulness for mental health specifically. Furthermore, more than four in five never-users reported little intention of using GenAI for mental health support in the future, which may partly explain the relatively stable prevalence pattern observed across the study year. Together, these findings suggest that current non-use often represents a principled and informed decision rather than unmet demand or digital exclusion. Since GenAI use for mental health is not universally acceptable preserving meaningful choice and maintaining access to high-quality human support remains essential.

Despite well-documented concerns regarding hallucinations, inappropriate advice and failures during safety-critical interactions, nearly three-quarters of users perceived their experiences with GenAI as beneficial, and perceived helpfulness was remarkably consistent across demographic and clinical subgroups. This is consistent with emerging Canadian evidence showing that most (84%) individuals who use AI for mental health support perceive it as helpful, despite comparatively low levels (17%) of public trust in these technologies^7^. Crucially, the impact findings should not be interpreted as evidence of clinical effectiveness or safety. Perceived benefit reflects users’ subjective appraisal rather than objective improvement in mental health outcomes. Thus, they should be interpreted alongside growing evidence that these systems remain vulnerable to factual errors, inappropriate reassurance and failures during high-risk mental health conversations. Rigorous prospective evaluations are therefore needed to determine when perceived benefit translates into measurable clinical benefit and when it may obscure potential harms, while also clarifying whether purpose-built GenAI mental health interventions offer safer and more effective approaches than general-purpose chatbots^22^.

This study has several strengths. To our knowledge, it is among the first population-based studies of GenAI use for mental health support in a Canadian university population and one of the few internationally to integrate prevalence, patterns of use, perceived impact, behavioural intentions, qualitative insights and multivariable correlates within a single theoretically informed framework. The WMH-ICS platform enabled linkage of GenAI use with validated psychiatric screeners and a broad range of psychosocial variables, while repeated cross-sectional sampling across an entire academic year provided a more robust picture of use than single time-point convenience surveys. Several limitations should also be considered. The cross-sectional design precludes conclusions regarding causality or temporal ordering, and all measures relied on self-report, introducing the potential for recall and reporting biases. Although the probability-based sampling strategy strengthens internal validity, the study was conducted at a single institution, which may limit generalisability to other institutions and settings. Finally, perceived helpfulness should not be interpreted as evidence of clinical efficacy, and future longitudinal and experimental studies are needed to determine the effectiveness, safety and longer-term consequences of GenAI use for mental health support.

In conclusion, general-purpose GenAI chatbots have rapidly become part of the informal mental health support landscape for university students. Use appears to be concentrated among students with greater psychological need, an existing orientation towards digital help-seeking and fewer interpersonal support resources, while many non-users consciously reject these technologies in favour of human connection. These findings suggest that GenAI should no longer be viewed as a speculative future technology within student mental healthcare but as an emerging component of contemporary help-seeking behaviour. The challenge for researchers, clinicians and universities is therefore how they can be evaluated, governed and integrated in ways that maximise potential benefit while safeguarding vulnerable users and preserving access to high-quality human care.

## Data Availability

All data produced in the present study are available upon reasonable request to the authors.

## Supplementary Data

**Supplementary Data Table 1:**
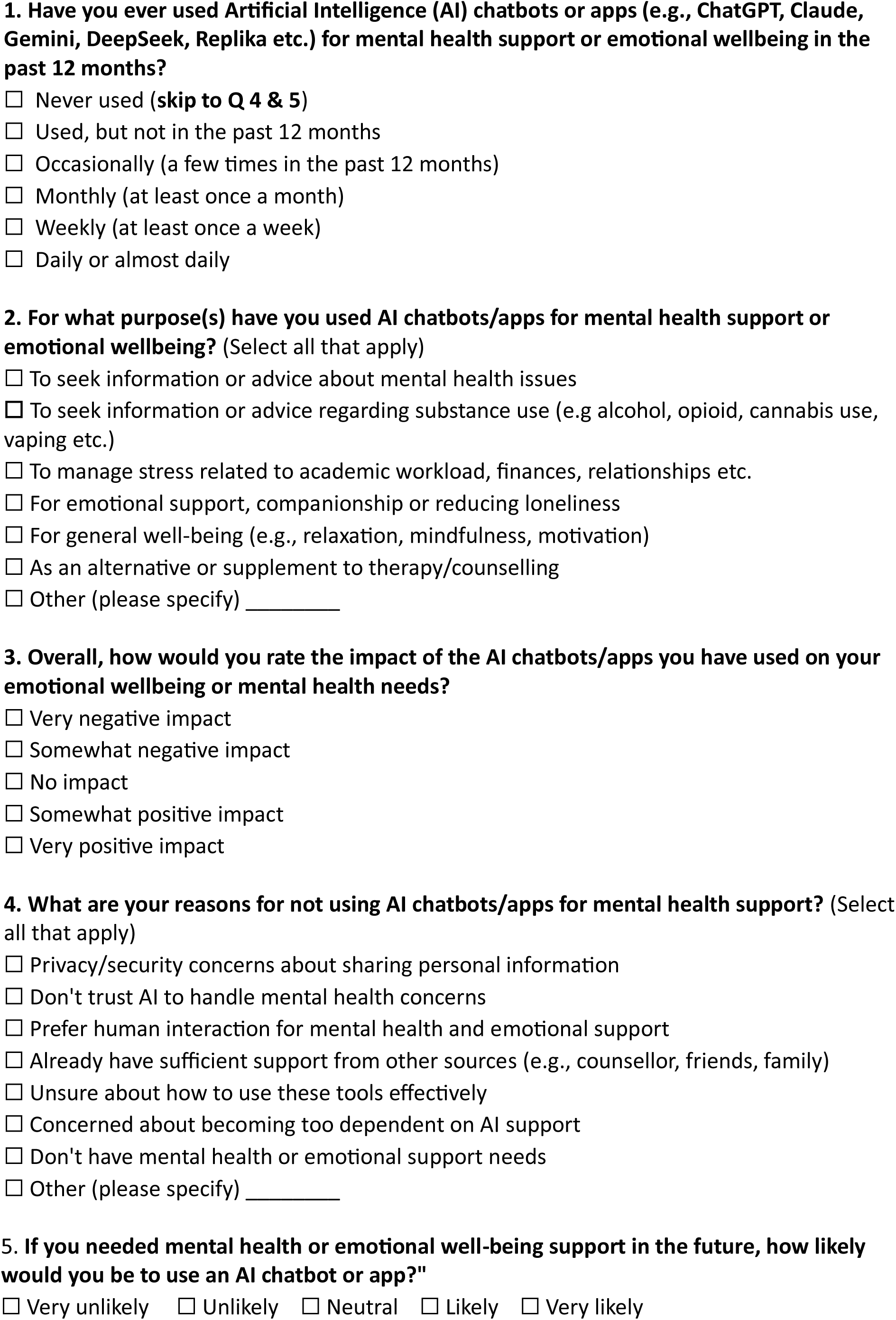
GenAI use related questions added to the WMH-ICS Survey.

**Supplementary Data Table 2:** Reflexive thematic analysis of free-text “Other” reasons for non-use. (n = 82 responses)

| Higher-order group | Theme | n | Illustrative quote(s) |
| --- | --- | --- | --- |
| Principled opposition to AI | General opposition/dislike of AI | 20 | "I boycott generative AI"; "Strongly against the use of generative AI" |
| Principled opposition to AI | Moral opposition | 6 | "AI is morally reprehensible to me"; "Generative AI steals other people's work" |
| Ethical & environmental | Environmental concerns | 16 | "Generative AI is bad for the environment"; "Ecological impact of AI usage" |
| Ethical & environmental | Ethical/environmental (combined) | 9 | "Ethical concerns of using AI (environmental, plagiarism, etc.)" |
| Distrust / perceived harm | Distrust AI for MH/harm concerns | 7 | "Giving personal health information to an AI chatbot is extremely dangerous... groups like me have been subject to discrimination"; "not appropriate for mental health treatment" |
| Distrust / perceived harm | Quality/accuracy doubts | 5 | "Gives inaccurate information"; "Not qualified to provide reliable mental health support" |
| Prefer human support | Prefer human/authentic interaction | 8 | "Why support AI when there are real people who went to school for their careers?" |
| Low salience / no need | Never considered/occurred | 8 | "It never occurred to me but I will"; "Never considered that as an option" |
| Low salience / no need | No need / support met elsewhere | 3 | "I have friends and family to talk to"; "I am an RN who has professional resources" |

**Supplementary Data Table 3:**
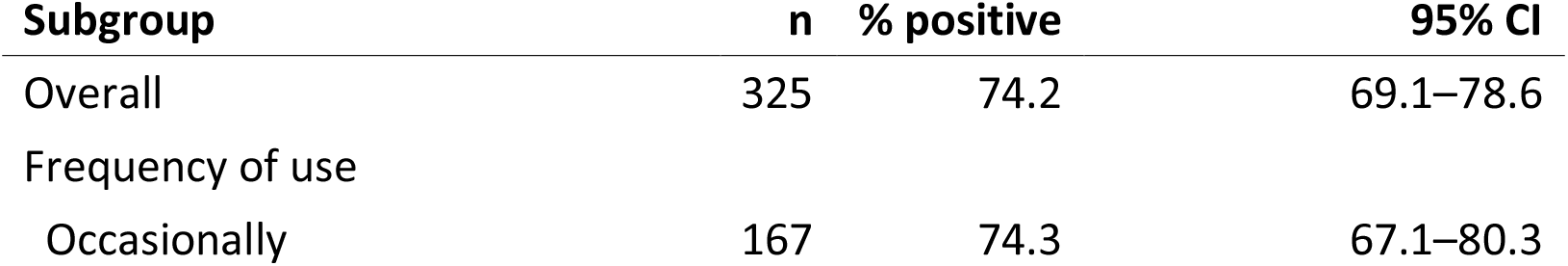
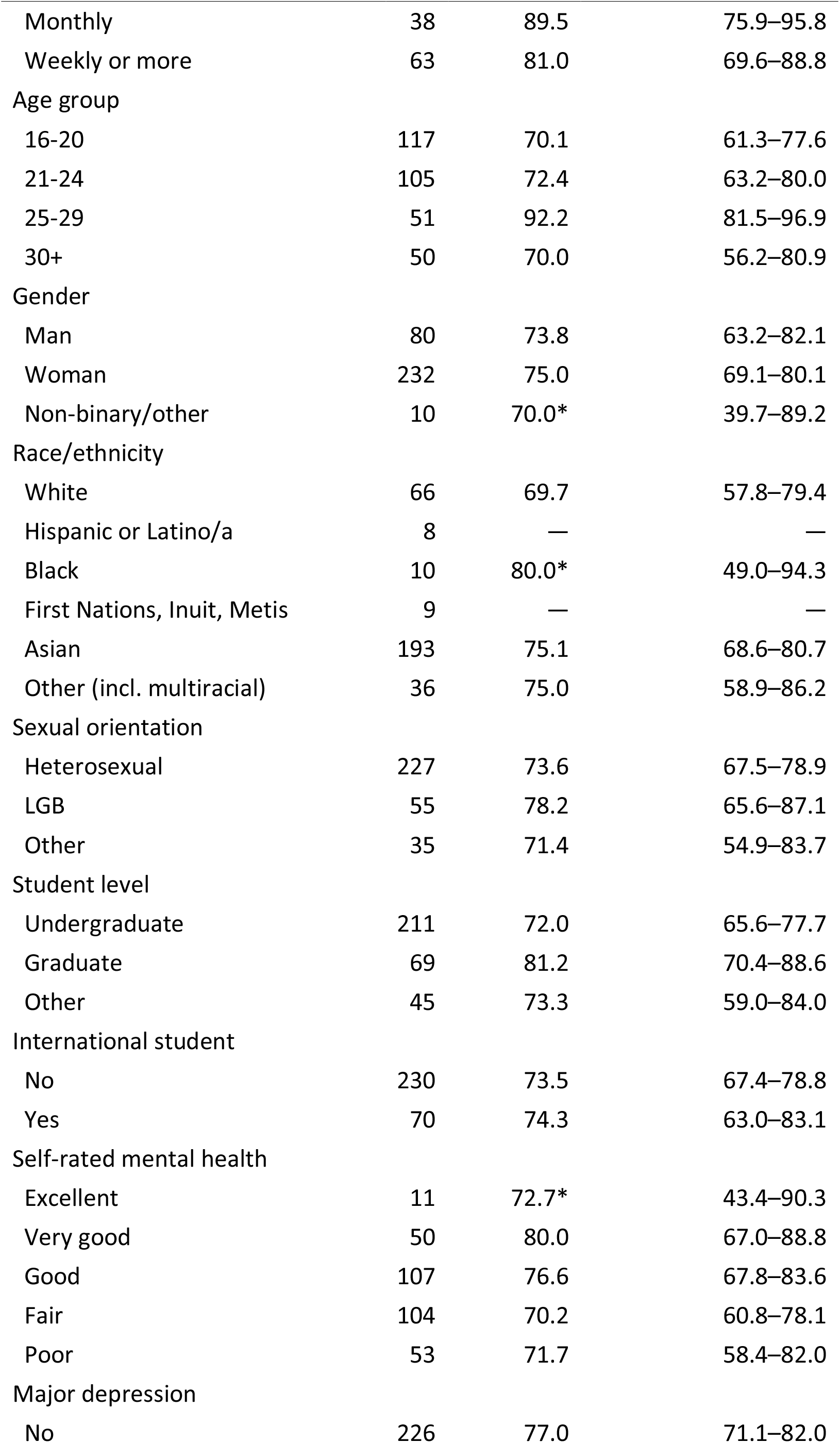
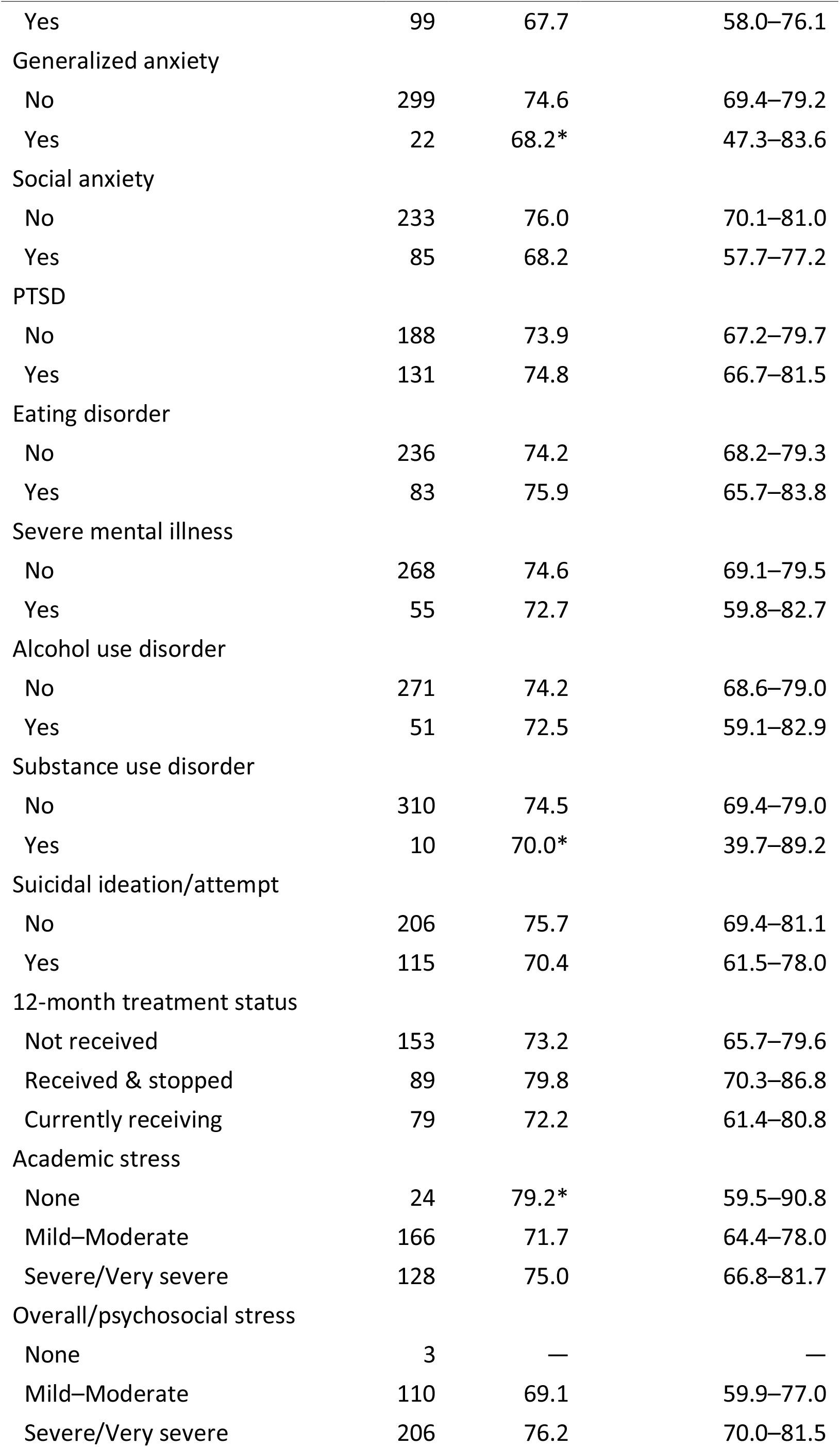
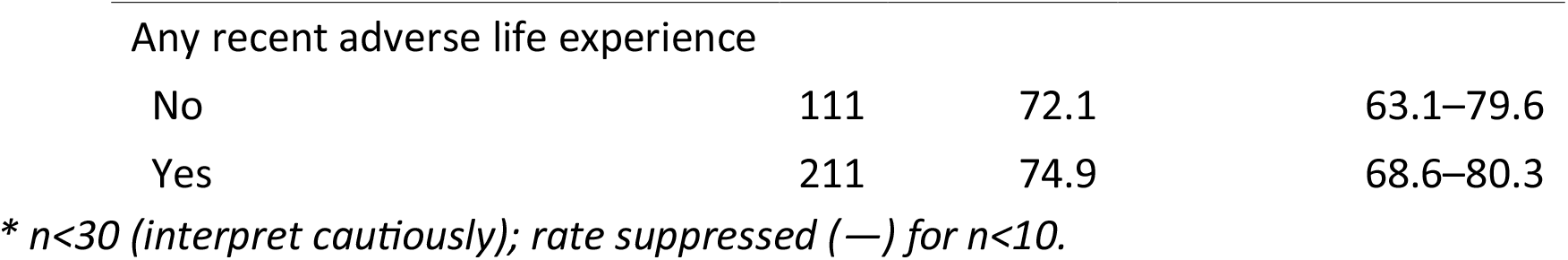
Perceived positive impact (somewhat/very positive) by subgroup among ever-users who rated impact. (n = 325).

| Subgroup | n | % positive | 95% CI |
| --- | --- | --- | --- |
| Overall | 325 | 74.2 | 69.1–78.6 |
| Frequency of use |  |  |  |
| Occasionally | 167 | 74.3 | 67.1–80.3 |

| **Subgroup** | **n** | **% positive** | **95% CI** |
| --- | --- | --- | --- |
| Monthly | 38 | 89.5 | 75.9–95.8 |
| Weekly or more | 63 | 81.0 | 69.6–88.8 |
| Age group |  |  |  |
| 16-20 | 117 | 70.1 | 61.3–77.6 |
| 21-24 | 105 | 72.4 | 63.2–80.0 |
| 25-29 | 51 | 92.2 | 81.5–96.9 |
| 30+ | 50 | 70.0 | 56.2–80.9 |
| Gender |  |  |  |
| Man | 80 | 73.8 | 63.2–82.1 |
| Woman | 232 | 75.0 | 69.1–80.1 |
| Non-binary/other | 10 | 70.0* | 39.7–89.2 |
| Race/ethnicity |  |  |  |
| White | 66 | 69.7 | 57.8–79.4 |
| Hispanic or Latino/a | 8 | — | — |
| Black | 10 | 80.0* | 49.0–94.3 |
| First Nations, Inuit, Metis | 9 | — | — |
| Asian | 193 | 75.1 | 68.6–80.7 |
| Other (incl. multiracial) | 36 | 75.0 | 58.9–86.2 |
| Sexual orientation |  |  |  |
| Heterosexual | 227 | 73.6 | 67.5–78.9 |
| LGB | 55 | 78.2 | 65.6–87.1 |
| Other | 35 | 71.4 | 54.9–83.7 |
| Student level |  |  |  |
| Undergraduate | 211 | 72.0 | 65.6–77.7 |
| Graduate | 69 | 81.2 | 70.4–88.6 |
| Other | 45 | 73.3 | 59.0–84.0 |
| International student |  |  |  |
| No | 230 | 73.5 | 67.4–78.8 |
| Yes | 70 | 74.3 | 63.0–83.1 |
| Self-rated mental health |  |  |  |
| Excellent | 11 | 72.7* | 43.4–90.3 |
| Very good | 50 | 80.0 | 67.0–88.8 |
| Good | 107 | 76.6 | 67.8–83.6 |
| Fair | 104 | 70.2 | 60.8–78.1 |
| Poor | 53 | 71.7 | 58.4–82.0 |
| Major depression |  |  |  |
| No | 226 | 77.0 | 71.1–82.0 |
| Yes | 99 | 67.7 | 58.0–76.1 |
| Generalized anxiety |  |  |  |
| No | 299 | 74.6 | 69.4–79.2 |
| Yes | 22 | 68.2* | 47.3–83.6 |
| Social anxiety |  |  |  |
| No | 233 | 76.0 | 70.1–81.0 |
| Yes | 85 | 68.2 | 57.7–77.2 |
| PTSD |  |  |  |
| No | 188 | 73.9 | 67.2–79.7 |
| Yes | 131 | 74.8 | 66.7–81.5 |
| Eating disorder |  |  |  |
| No | 236 | 74.2 | 68.2–79.3 |
| Yes | 83 | 75.9 | 65.7–83.8 |
| Severe mental illness |  |  |  |
| No | 268 | 74.6 | 69.1–79.5 |
| Yes | 55 | 72.7 | 59.8–82.7 |
| Alcohol use disorder |  |  |  |
| No | 271 | 74.2 | 68.6–79.0 |
| Yes | 51 | 72.5 | 59.1–82.9 |
| Substance use disorder |  |  |  |
| No | 310 | 74.5 | 69.4–79.0 |
| Yes | 10 | 70.0* | 39.7–89.2 |
| Suicidal ideation/attempt |  |  |  |
| No | 206 | 75.7 | 69.4–81.1 |
| Yes | 115 | 70.4 | 61.5–78.0 |
| 12-month treatment status |  |  |  |
| Not received | 153 | 73.2 | 65.7–79.6 |
| Received & stopped | 89 | 79.8 | 70.3–86.8 |
| Currently receiving | 79 | 72.2 | 61.4–80.8 |
| Academic stress |  |  |  |
| None | 24 | 79.2* | 59.5–90.8 |
| Mild–Moderate | 166 | 71.7 | 64.4–78.0 |
| Severe/Very severe | 128 | 75.0 | 66.8–81.7 |
| Overall/psychosocial stress |  |  |  |
| None | 3 | — | — |
| Mild–Moderate | 110 | 69.1 | 59.9–77.0 |
| Severe/Very severe | 206 | 76.2 | 70.0–81.5 |
| Any recent adverse life experience |  |  |  |
| No | 111 | 72.1 | 63.1–79.6 |
| Yes | 211 | 74.9 | 68.6–80.3 |

\* $n < 30$ (interpret cautiously); rate suppressed (—) for $n < 10$ .

**Supplementary Data Table 4:**
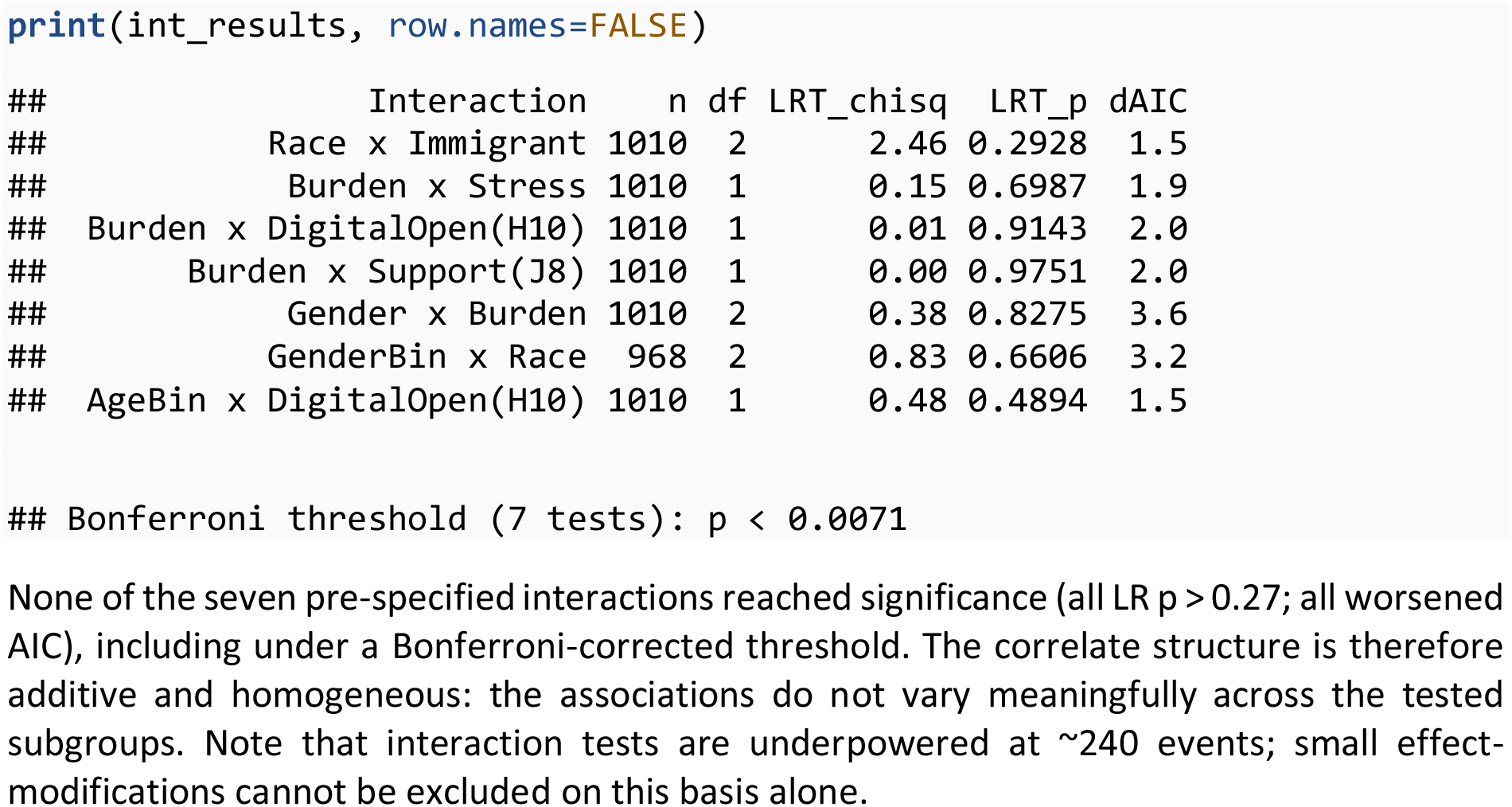
Pre-specified interactions. We tested a pre-specified family of seven interactions, in two tiers: primary (theory-driven) — race × immigrant, clinical-burden × stress, clinical-burden × digital-openness, clinical-burden × social-support; and secondary (effect-modification checks) — gender × burden, gender × race (gender binarised for cell adequacy), age × digital-openness.

**Supplementary Data Table 5:**
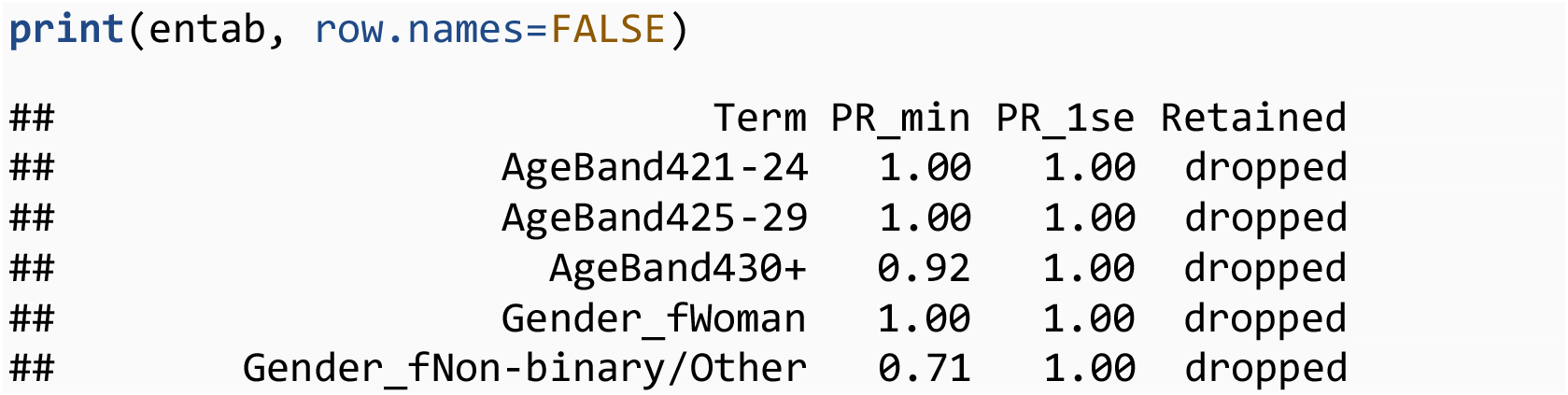
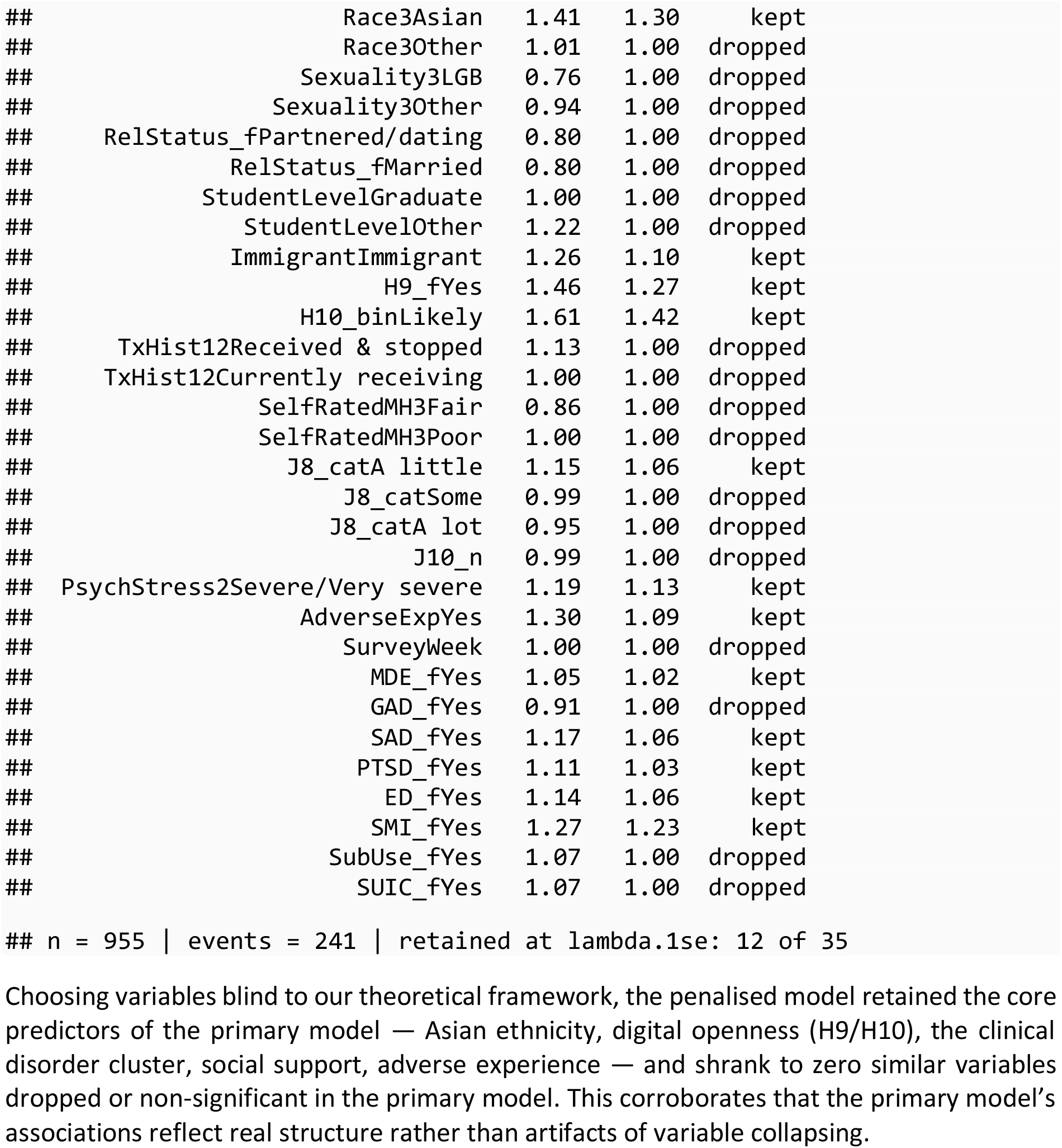
Elastic Net Sensitivity Analysis. To confirm results and to verify that variable modification e.g., collapsing clinical indicators did not manufacture the findings, we fit an **elastic-net penalised Poisson model** (alpha = 0.5) on the *full uncollapsed* predictor set — all individual disorders, stress, self-rated health, network size, and un-collapsed factors restored.

